# Prevalence and correlates of Metabolic Syndrome among adults in Freetown, Sierra Leone: A comparative analysis of NCEP ATP III, IDF and Harmonized ATP III criteria

**DOI:** 10.1101/2023.06.26.23291915

**Authors:** James Baligeh Walter Russell, Theresa Ruba Koroma, Santigie Sesay, Sallieu K Samura, Sulaiman Lakoh, Ansumana Bockarie, Onomeh Thomas Abiri, Victor Conteh, Sorie Conteh, Mohamed Smith, O Z Mahdi, Durodami. R. Lisk

## Abstract

**Background:** Metabolic syndrome (MS) is a global health concern, especially for low and middle-income countries with limited resources and information. The study’s objective was to assess the prevalence of MS in Freetown, Sierra Leone, using the Adult Treatment Panel III (ATP III), International Diabetes Federation (IDF) and Harmonize ATP III. Additionally, we aimed to establish the concordance between these three different criteria used.

**Methods:** This community-based health screening survey was conducted from October 2019 to October 2022. A multistage stratified random design was used to select adults aged 20 years and above. Mean, interquartile range (IQR), and logistic regression were used for statistical analysis. The kappa coefficient statistics resolved the agreement between these defined criteria.

**Results:** The prevalence for NCEP ATP III, Harmonize ATP III and IDF criteria was 11.8% (95% CI: 9.0 - 15.15), 14.3% (95% CI: 11.3 - 18.0), and 8.5% (95% CI: 6.2 - 11.2), respectively for the 2,394 selected adults. The kappa coefficient (κ) agreement between the MS is: Harmonized ATP III and IDF criteria = [(208 (60.8%); (κ =0.62)]; Harmonized ATP III and NCEP ATP III = [(201 (58.7%); (κ =0.71)]; while IDF and NCEP ATP III was [(132 (38.6%); (κ =0.52)]. In the multivariable regression analysis, waist circumference correlated with all three MS criteria: ATP III [AOR = 0.85; C.I 95%: (0.40-1.78), p = 0.032], Harmonized ATP III [AOR = 1.14; C.I 95%: (0.62-2.11), p = 0.024], IDF [AOR = 1.06; C.I 95% (0.52-2.16), p = 0.018]

**Conclusion:** We reported a high prevalence of MS in Freetown, Sierra Leone and identified waist circumference as a major risk factor for MS. This underscores the crucial role of health education and effective management of MS in Sierra Leone.

## 1. Introduction

Metabolic syndrome (MS) is a group of closely related risk factors associated with an increased risk of cardiovascular diseases and type 2 diabetes [1–3]. Studies have shown that people with MS are three times more likely to suffer a stroke or heart attack than those without MetS [4,5]. The risk factors associated with MetS include elevated blood pressure, high triglycerides, low high-density lipoprotein cholesterol (HDL-C), high fasting glucose, and central obesity [6]. Early detection and proper treatment of MS are crucial to preventing serious health conditions, as they are essential for optimal health outcomes. Therefore, it is important to prioritize these measures to prevent potential health risks. [7]. Leading public health experts and professional organizations have validated different diagnostic criteria for MS [8–13]. Each criterion has recommended cutoff values that are distinct and essential for diagnosing MS. To combine the various criteria of these organizations, Alberti et al proposed the Harmonized National Cholesterol Education Program (NCEP) Adult Treatment Panel (ATP) III criteria. [14]. Nevertheless, the accuracy of MetS criteria in predicting cardiovascular diseases is still controversial. Koutsovasilis et al. suggested that the International Diabetes Federation (IDF) criterion better predicts acute coronary syndrome in NCEP ATP III than harmonised ATP III, but in a similar study, Nilsson did not find evidence supporting IDF’s superiority [15,16]. Further research is therefore needed to understand this issue.

In several Sub-Saharan African (SSA) countries, the prevalence of non-communicable diseases (NCDs) has increased because of the shift from communicable diseases. This change can be attributed to increased urbanization and changing lifestyles in many low- and middle-income countries. [17]. Hence, it can be inferred that metabolic syndrome is common, given the high prevalence of cardiovascular diseases and diabetes in SSA.[1]. However, there is a lack of information on MetS in the SSA region, as most research on MetS is conducted in North America, Europe, and Asia. [18,19]. Notwithstanding, most data on MetsS in Sub-Saharan Africa emanate from small clinical studies, with a limited number of documented reports on epidemiological studies. [20].

The lack of epidemiological data on MS in Sierra Leone must be addressed urgently to determine the appropriate interventions required to mitigate its effects on the population. Our study aimed to determine the prevalence of MS among adult Sierra Leoneans by using three different definitions: the ATP III, IDF, and Harmonized ATP III Criteria. Furthermore, we sought to evaluate the level of agreement between the three diagnostic criteria for metabolic syndrome.

## 2. METHODS

### 2.1. Study area, study design, and sampling technique

This community health-screening survey was conducted in Freetown, Western Area Urban, Sierra Leone, between October 2019 and October 2022. A multistage stratified random sampling design consisting of four stages was used to select adults aged 20 years and above. The details of the methods applied in the “Ecobank Health Screening and Awareness Programme” in Sierra Leone are found in our previous study [21].

### 2.2. Socio-demographic, behavioural and clinical characteristics, and anthropometric and blood pressure measurements

Socio-demographic information such as age, sex, gender, educational level, religion, marital status, smoking status, and alcohol consumption was collected. We also collected information on the medical history (including a family history of hypertension and diabetes mellitus), fruit and vegetable consumption, and physical activity. The details of the blood pressure and arthrometric (height, body weight and waist circumference) measurements are found in our previous study [21].

### 2.3. Inclusion and Exclusion Criteria

The individuals included in the study were adults who were 20 years of age or older and had lived in the city for a minimum of twelve months. Pregnant and lactating mothers, participants with mental illness/dementia and persons unwilling to grant consent were excluded from the study.

### 2.4. Clinical biochemistry measurements

Blood sample (3ml) was taken from each consented participant to estimate biochemical parameters such as fasting glucose, total cholesterol, triglycerides, HDL-C, and low-density lipoprotein cholesterol (LDL-C) [21].

#### Criteria for MS

**Table 1.** Diagnostic criteria and cutoff points of metabolic syndrome according to various organizations.

| Parameter/criteria | Harmonized criteria -2009 | IDF-2005 | NCEP ATP III-2001 |
| --- | --- | --- | --- |
| Prerequisites for diagnosing MetS | Any three of the following | Abdominal obesity along with any other two | Any three of the following |
| Waist Circumference (cm) |  |  |  |
| Men | ≥ 90 cm | ≥ 90 cm | ≥102 cm |
| Woman | ≥ 80 cm | ≥ 80 cm | ≥ 88 cm |
| HDL-C |  |  |  |
| Men < 40 | < 1.03 mmol/L | < 1.03 mmol/L | < 1.03 mmol/L |
| Woman <50 | < 1.30 mmol/L | < 1.30 mmol/L | < 1.30 mmol/L |
| Blood pressure (mmHg) | 130/85mmHg | 130/85mmHg | 130/85mmHg |
| Fasting | 5.6 mmol/l | 5.6 mmol/l | 6.1 mmol/l |
| Triglyceride | ≥1.70 mmol/L (150 mg/dL) | ≥1.70 mmol/L (150 mg/dL) | ≥1.70 mmol/L (150 mg/dL) |

### 2.5. Ethical approval and registration

The Sierra Leone Ethics and Scientific Review Committee approved the research protocol, questionnaire, and consent form. The study protocol was registered with the Research Registry and assigned the unique identification number researchregistry8201 [https://www.researchregistry.com/browse-the-registry#home/ The credibility and reliability of our methods and findings strictly adhered to the guidelines outlined in the STROBE statement. [22].

### 2.6. Statistical Analyses

IBM SPSS Statistical 2.6 and STATA 17 software were used to analyze the dataset of this study. All baseline characteristics of metabolic syndrome were examined. Median and IQR were used as necessary. Univariate and multivariable logistic regression was conducted to explore the relationship between demographic characteristics and cardiovascular risk factors. We included several independent variables such as age, BMI, waist circumference, fruit and vegetable intake, blood pressure, alcohol consumption, smoking status, diabetes mellitus, total cholesterol, HDL-C, LDL-C, triglycerides, and physical activity levels. A two-tailed p-value of ≤ 0.05 was considered statistically significant. The Kappa statistic was used to evaluate the level of agreement among three criteria. NCEP ATP III, Harmonized ATP III, and IDF criteria were used to calculate the prevalence of each component of MetS.

## 3. RESULTS

### 3.1. Basic characteristics of the study

Of the 2,394 participants recruited for this study, 1250 (52.2%) were male. The median age of the participants was 41.6 years [IQR:34 - 49] years. Age, height, waist circumference, and total cholesterol levels showed statistical differences, as shown in Table 2.

**Table 2:**
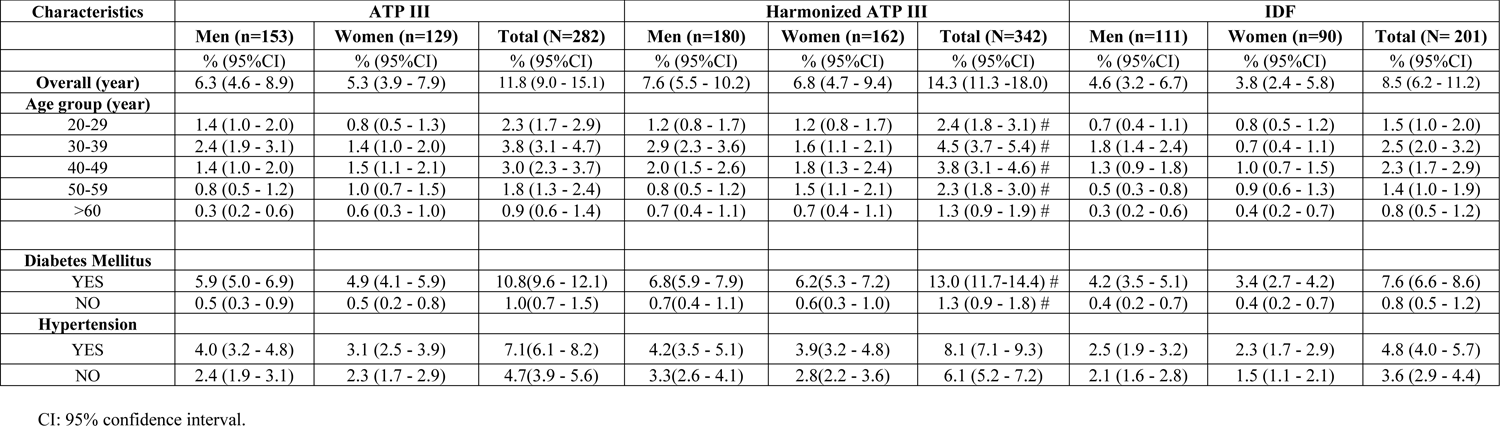
Prevalence of metabolic syndrome among study population

### 3.2. Prevalence of metabolic syndrome

The MS prevalence rates for NCEP ATP III, Harmonized ATP II criteria and IDF criteria were 11.8% [95% CI: (9.0 - 15.2)], 14.3% [95% CI: (11.3 - 18.0)], and 8.5% [95% CI: (6.2 - 11.2)], respectively (Table 3). Using the ATP III, Harmonized ATP III, and IDF criteria, the prevalence of MS for men was 6.3% (95% CI: 4.6 - 8.9), 7.6% (95% CI: 5.5 - 10.2), and 4.6% (95% CI: 3.2 - 7.9), respectively. For women, the prevalence rates were 5.3% (95% CI: 3.9 - 21.1), 6.8% (95% CI: 4.7 - 9.4), and 3.8% (95% CI: 2.4 - 5.8), respectively. For men, the highest prevalence of MS according to ATP III [2.4% (95% CI: 1.9 - 3.1)], Harmonized ATP III [2.9% (95% CI: 2.3 - 3.1)], and IDF criteria [1.8 % (95% CI: 1.4 - 2.4)] were documented in the age group 30 - 39 years. Unlike women, the highest sex-standardized prevalence for ATP III [1.5% (95% CI: 1.1 - 2.1)], Harmonized ATP III [1.8% (95% CI: 1.3 - 2.4)], and IDF criteria [1.0% (95% CI: 0.7 - 1.5) were recorded in a much older age group 40 - 49 years. The prevalence of Metabolic Syndrome among diabetic and hypertensive individuals is also represented in Table 2.

**Table 3.** Demographic, anthropometric and biochemical variables of study population.

| Characteristics | Total (2394) | Male (1250) | Female (1144) | p-value |
| --- | --- | --- | --- | --- |
|  | Median(IQR) | Median(IQR) | Median(IQR) |  |
| Age | 41.6 (34-49) | 39(33-48) | 42(34-49) | **0.043** |
| Weight | 70.4(61.8-85.4) | 69.9(61.6-85.1) | 70.5(61.7-85.0) | 0.634 |
| Height | 1.73(1.70-1.77) | 1.74(1.70-1.77) | 1.73(1.70-1.77) | **0.012** |
| BMI | 23.7(21.1-27.8) | 23.6(20.9-27.6) | 23.8(20.9-27.7) | 0.107 |
| WC | 89.0(78.7-92.7) | 92.6(89.8-96.7) | 78.8(75.9-82.7) | **<0.001** |
| SBP | 126(107-148) | 127(106-146) | 125(106-147) | 0.178 |
| DBP | 85(77-94) | 86(78-94) | 85(75-94) | 0.126 |
| Triglyceride | 1.53(1.45-1.63) | 1.53(1.45-1.62) | 1.54(1.45-1.65) | 0.447 |
| Total Cholesterol | 4.79(4.67-5.00) | 4.65(4.67-4.89) | 4.91(4.66-5.90) | **<0.001** |
| Low HDL-C | 1.34(1.26-1.41) | 1.34(1.26-1.41) | 1.34(1.25-1.41) | 0.532 |
| LDL-C | 2.97(2.56-3.20) | 2.99(2.54-3.21) | 2.95(2.51-3.17) | 0.118 |
| FBS | 4.70(4.10-5.40) | 4.80(4.10-5.40) | 4.70(4.10-5.40) | 0.249 |
| HBA1C | 5.10(4.60-5.60) | 5.10(4.60-5.60) | 5.10(4.8-5.6) | 0.481 |

**Table 4.** Agreement among the different criteria in diagnosis of metabolic syndrome.

| Harmonised criteria |  |  |  |  |  |
| --- | --- | --- | --- | --- | --- |
|  |  | MetS + | MetS- | Total | Kappa (95% CI) |
| IDF | MetS+ | 208 | 74 | 282 | 0.62(0.59 - 0.64) |
|  | MetS- | 134 | 1978 | 2112 |  |
|  | Total | 342 | 2051 | 2394 |  |
| Harmonised criteria |  |  |  |  |  |
| ATP III | Met + | 201 | 0 | 201 | 0.71(0.69 - 0.73) |
|  | Met - | 141 | 2052 | 2193 |  |
|  | Total | 342 | 2052 | 2394 |  |
| NCEP ATP III |  |  |  |  |  |
| IDF | Met + | 132 | 0 | 132 | 0.52(0.50 - 0.54) |
|  | Met - | 210 | 2052 | 2262 |  |
|  | Total | 342 | 2052 | 2394 |  |

### 3.3. Agreement among NCEP ATP III, Harmonized ATP III, and IDF criteria

The age-standardized distribution for most MS criteria by sex and age group is similar, except for waist circumference criteria (Figure 1). Of all the participants with metabolic syndrome, the Venn diagram depicted 8.6% as having MS by all three criteria. (Figure 2). Agreement between Harmonized ATP III and IDF criteria was 208 (60.8%) with kappa statistics of 0.62, while the agreement between Harmonized ATP III and NCEP ATP III was 201 (58.7%) with kappa statistics of 0.71. All participants diagnosed with MS by IDF also met the criteria of Harmonized ATP III (k=0.52). The highest agreement was observed between Harmonized ATP III and NCEP ATP III (k = 0.71), while the lowest was observed between NCEPATP III and IDF criteria (k = 0.52).

**Fig 1.**
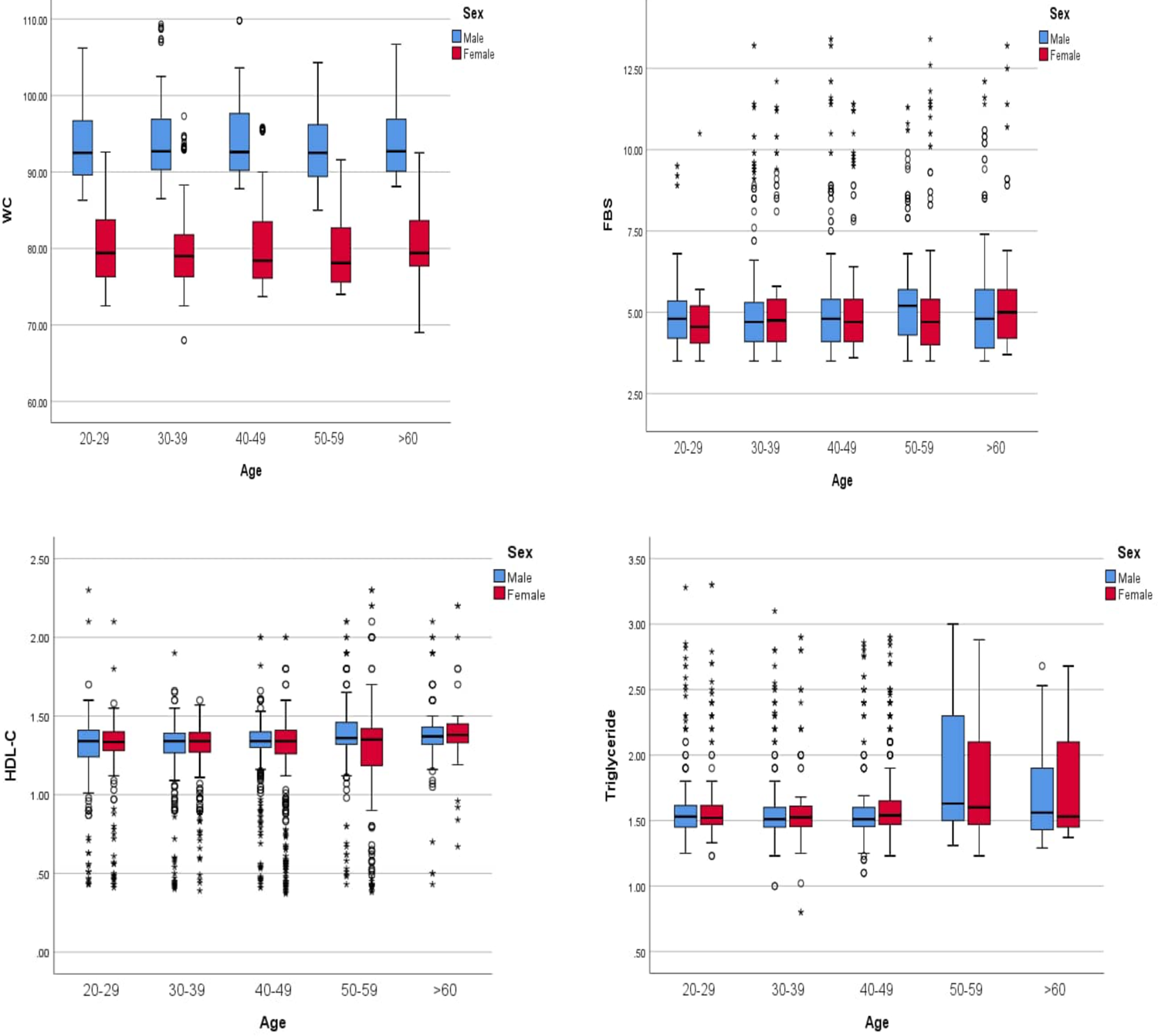
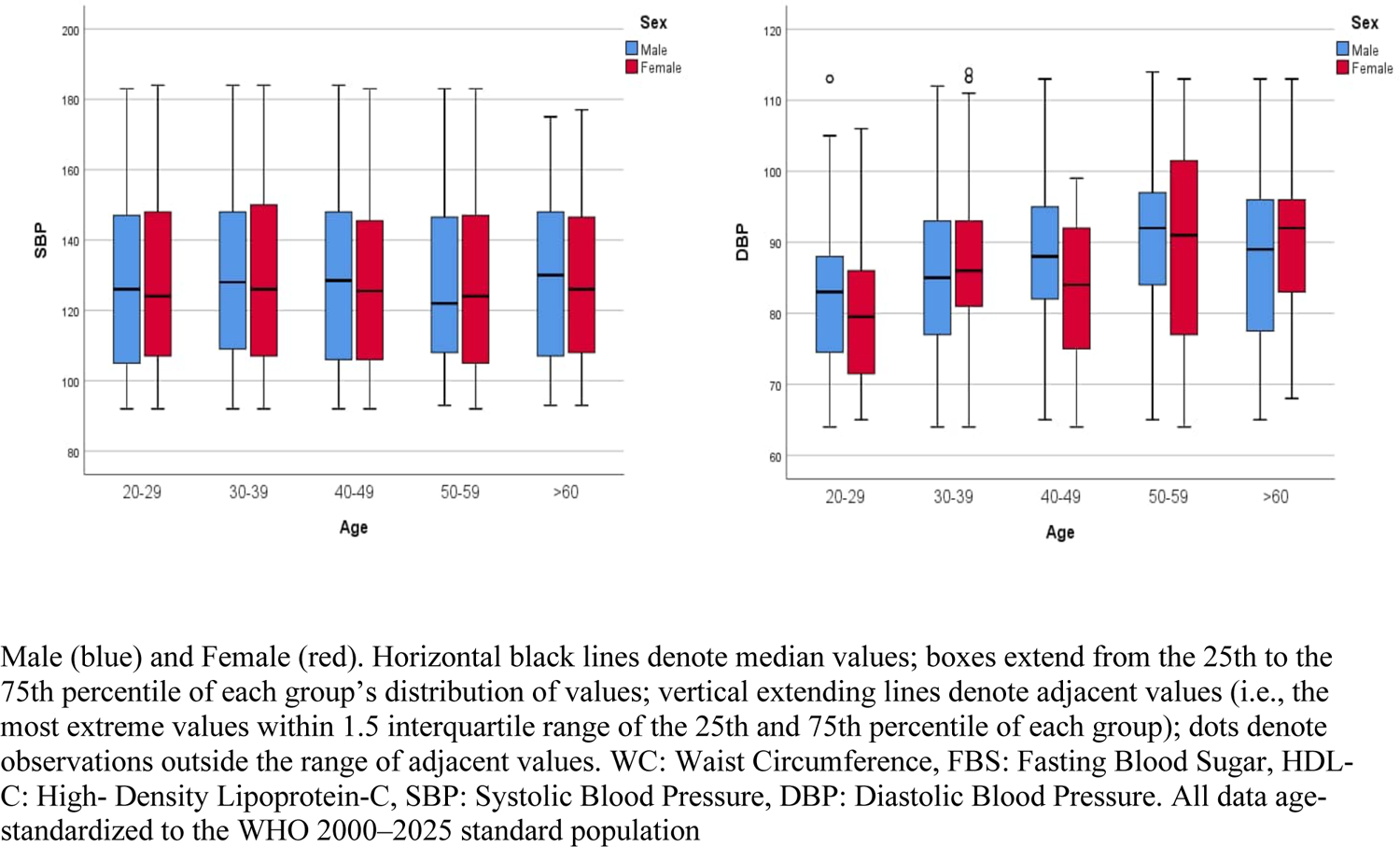
Boxplot showing comparative analysis of the different risk factors of MetS by age and sex.

**Fig 2.**
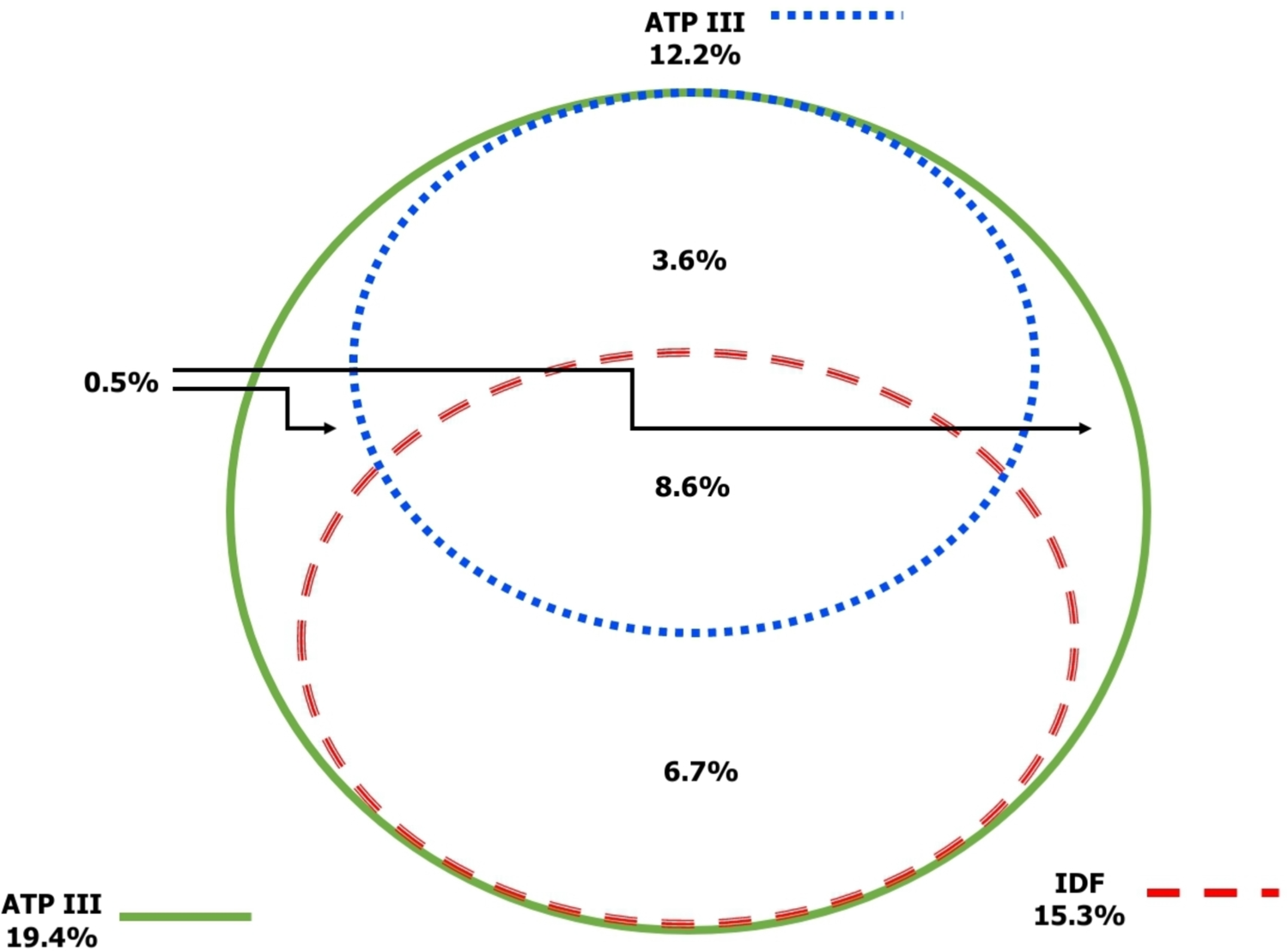
Overall crude prevalence of MetS according to ATP III, IDF and Harmonized ATP III criteria.

### 3.4. Logistic Regression Model among NCEP ATP III, Harmonized ATP III, and IDF criteria

We included several independent variables such as age, BMI, waist circumference, fruit and vegetable intake, blood pressure, alcohol consumption, smoking status, diabetes mellitus, total cholesterol, HDL-C, LDL-C, triglycerides, and physical activity levels in the stepwise logistic regression model to analyse the various MS criteria (Table 4).

**Table 4.**
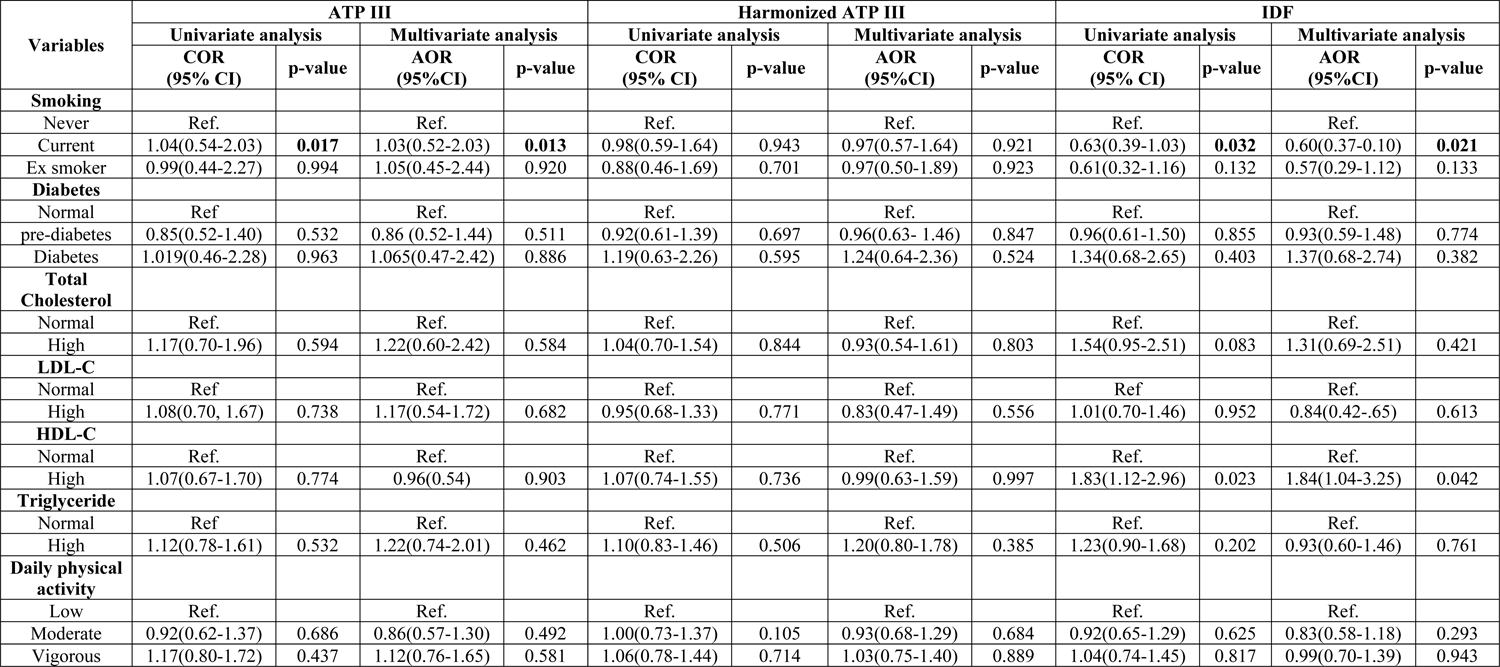
continuation: Logistic regression analysis of risk factors for metabolic syndrome.

**Table 5.**
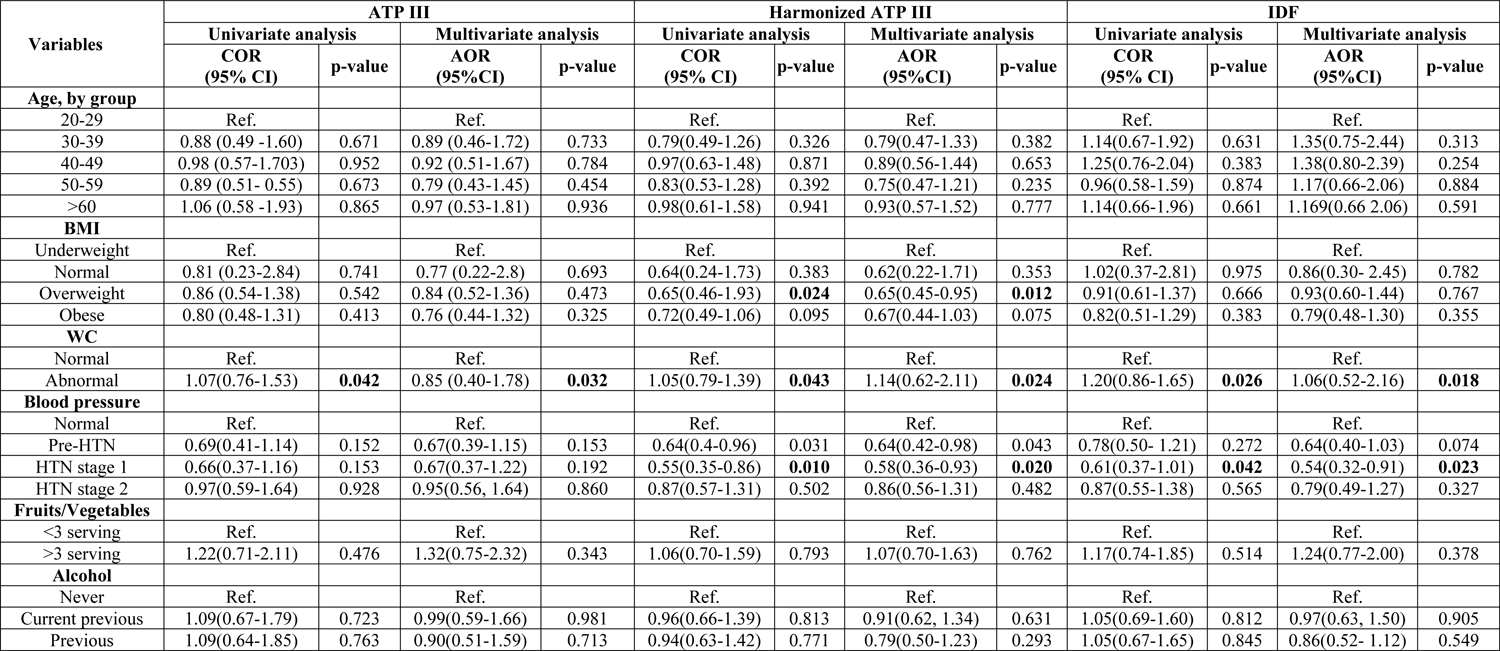
Logistic regression analysis of independent variables for metabolic syndrome.

WC as a risk factor correlated with all the MS criteria: NCEP ATP III, Harmonized ATP III, and IDF. In the NCEP ATP III model, the crude odds ratio was 1.07 with a 95% confidence interval of (0.76-1.5) and a p-value of 0.04. After adjusting for relevant factors, the adjusted odds ratio was 0.85 [95% CI (0.40-1.78)], p = 0.036. Similarly, in the Harmonized ATP III model, the crude odds ratio was 1.05 with a 95% confidence interval of (0.79-1.39) and a p-value of 0.045. The adjusted odds ratio was 1.14 with a 95% confidence interval of (0.62 - 2.11) and a p-value of 0.02. In the IDF model, the crude odds ratio was 1.20 with a 95% confidence interval of (0.86-1.65) and a p-value of 0.026, while the odds ratio was 1.06 with a 95% confidence interval of (0.52-2.16) and a p-value of 0.018.

Stage 1 Hypertension is associated with both ATP III and IDF. For NCEP ATP III, the Crude OR is 0.55 and the C.I is 95% (0.35-0.86) with a p-value of 0.01, while the adjusted OR is 0.58 with a C.I of 95% (0.36-0.93) and a p-value of 0.04. The preliminary odds ratio in the IDF is 0.6, with a confidence interval of 95% (0.37-1.01) and a p-value of 0.44. However, after adjustments, the odds ratio is 0.54 with a confidence interval of 95% (0.32-0.91) and a p-value of 0.02. Smoking is a risk factor that was found to be associated with NCEP ATP III and IDF. The data showed that for NCEP ATP III, the crude odds ratio was 1.04 with a 95% confidence interval of (0.54 - 2.03) and a p-value of 0.01, while the adjusted odds ratio was 1.03 with a 95% confidence interval of (0.52-2.03) and a p-value of 0.01. For IDF, the crude odds ratio was 0.63 with a 95% confidence interval of (0.39-1.03) and a p-value of 0.04, while the adjusted odds ratio was 0.60 with a 95% confidence interval of (0.37-0.10) and a p-value of 0.05.

Several variables, including age, fruit and alcohol consumption, diabetes mellitus, total cholesterol, HDL-C, LDL-C, triglyceride levels, and daily physical activities, did not demonstrate an independent association with Metabolic Syndrome (MetS) when other covariates were considered.

## 4. Discussion

This community-based health screening survey for NCD has provided extensive data on MetS. It is the first survey to document the prevalence of MetS in Sierra Leone based on three different criteria (NCEP ATP III, Harmonized ATP III, and IDF) and the second-largest dataset on MetS in the West African subregion, following Nwamko et al [23]. The current study found that the prevalence of Metabolic Syndrome (MetS) varied according to the diagnostic criteria used.

In recent years, metabolic syndrome has become a common disorder worldwide, with different African regions experiencing varying prevalence rates [20]. Factors such as urbanization, industrialization, an ageing population, criteria used to define MetS, sampling methods, lifestyle choices, and genetic variations, are believed to contribute to this trend [17]. The prevalence rate for each MetS criterion in our study population was 11.8% for NCEP ATP III, 14.3% for Harmonized ATP III criteria and 8.5% for IDF criteria. Using the NCEP ATP III criterion, the prevalence of MetS was lower than reported in Khartoum, Sudan (19.8%), Assin Fosu, Ghana (37.1%), Ogbomoso, Nigeria (33.0%), Ilara-Akaka, Nigeria (21.1%), Dakar, Senegal (15.7%), and Cape Town, South Africa (55.4%), but relatively higher than reported in Brazzaville, Congo (8.7%), Abuja, Nigeria (8.8%) and Mizan-Aman, Ethiopia (9.6%) [24–32]. However, our study found that the prevalence of Harmonised ATP III was comparable to the reported 15.1% from China, but lower than the rates of 23.6% in Nigeria and 40.7% in Ethiopia. [33–35]. Using IDF, the prevalence of MS was similar to the reported rates in Cotonou, Benin (7.4%) and Bondo District, Kenya (8.5%), but lower than rates found in Ghana, South Africa, and Nigeria. [36–40].

Recent data from a systematic review and meta-analysis on the prevalence of MetS in SSA according to the different diagnostic criteria are consistent with some of our findings [20]. Our study demonstrates that metabolic syndrome is a serious public health challenge in Sierra Leone. Like other African countries, Metabolic syndrome is becoming more common and should be given sufficient attention.

Males are at a greater risk for developing Metabolic Syndrome (MetS) than females in this study. Using all the three defined criteria, the prevalence of MetS is higher in men than women, and this is similar studies reported in Abuja, Nigeria; Brazzaville, Congo; Nairobi, Kenya; Mwanza, Tanzania; Cotonou, Benin; Khartoum, Sudan; Northern province, Ethiopia; North-west province, South Africa; North-West, Nigeria and Yaoundé, Cameroon [41–45]. It is absolutely crucial that cardiovascular diseases related to Metabolic Syndrome (MetS) are prevented and effectively managed, especially in men. This must be the top priority in any healthcare strategy to optimise patient outcomes.

We investigated the prevalence of MetS by age group and found that the highest prevalence was observed in the third decade for both males and females for all three defined criteria. The high prevalence rate of MetS in the third decade is due to the youthful cohort of our study, as almost half of our population is under 40 years. Ageing as an independent cardiovascular risk is associated with the evolution of insulin resistance and accumulation of visceral adipose tissue, which is important in the pathogenesis of MetS [46,47]. Our analysis to identify the prevalence of MetS amongst diabetic individuals indicated a lower prevalence than studies reported from Africa [48–50]. Whereas most African research on Mets and diabetics are most often hospital-based studies, our findings reflect a community-based screening in metabolic syndrome, which may account for the lower prevalence. Since hypertension is a major component of MetS and a widely recognised risk factor for cardiovascular disease, we evaluated the prevalence of MetS among individuals with hypertension. Consistent with other research conducted in Africa, our findings suggest that hypertension contributes to the exacerbation of MetS [51–53].

The threshold for WC among Black Africans is still controversial as there is no robust research related to a validated cut-off. However, most African studies use thresholds of 102cm for men and 88cm for women to define abdominal obesity among African indigenes [19,54]. Due to urbanization and lifestyle changes, many African countries are experiencing increased abdominal obesity. In our studies, the waist circumference distribution by sex and group was significantly different when adjusted for age, with men having a higher WC distribution. The high cut-off for men may explain the disparity in WC distribution by sex and age group in our study, which is consistent with other African studies [5,13,17,27]. The IDF and Harmonized ATP III criteria use lower cut-off values than the NCEP ATP III criteria and may lead to the inclusion of individuals with lower levels of these risk factors. The mandatory use of a low cut-off of WC (male > 90cm, females > 80cm) and any other risk factors for IDF definition may account for the low prevalence of MetS in our study.

Therefore, some individuals with MetS may not be identified using the IDF criteria. On the other hand, the Harmonised ATP III definition is non-discriminatory and requires a minimum of any three risk factors for its diagnosis. As a result, the probability of having more individuals being diagnosed with MeS is high. This may be the reason for the high prevalence of MetS in Harmonised ATP III definition in our study.

Our study showed that Harmonised ATP III and IDF criteria gave better agreement, while the agreement between NCEP ATP III and IDF was good. However, the agreement between Harmonised ATP III and NCEP ATP III criteria appeared to be the best. The significant overlap among the MetS criteria in our study is not unexpected, as other studies have also reported similar findings in Nigeria, Brazil, India and Mongolia [34,55–57]. Even though MetS has been extensively researched in the last decade, public awareness is limited in most LMIC and industrial countries. Our study, therefore, showed that MetS is a public health issue in Sierra Leone, requiring much-needed public awareness.

Our study’s regression analysis of demographics, anthropometrics, lifestyle, biochemical factors, and MetS are consistent with previous African literature [17,24,29,43]. On logistic regression analysis, WC was the only variable correlated with all three defined MetS criteria using both unadjusted and adjusted ratios. The relationship between WC and obesity health-related risks can be attributed to factors such as low physical activity and high-energy diets. The relationship underscores the existence of many abnormalities in people with metabolic syndrome. Current smoking was found to be associated with both the unadjusted and adjusted ratio in the ATP III and IDF criteria, which was unlikely in the NCEP ATP III criterion.

There is a clear correlation between smoking and MetS, which is supported by increased circulating hormones like cortisol, catecholamines, vasopressin and growth hormones [58]. As a causal factor in the development of MetS, it is therefore imperative that individuals should understand the associated risk of smoking and take steps to quit or avoid smoking. When unadjusted and adjusted ratios were analysed, there was a correlation between stage 1 hypertension and the MetS criteria for Harmonized ATP III and IDF criteria. Since hypertension is a major cause of metabolic syndrome, it predisposes individuals to the risk of developing cardiovascular diseases. [51]. Hence blood pressure control is imperative in preventing many cardiovascular morbidity and mortality.

### 4.1. Limitations and strengths of the study

When interpreting our study, it is important to consider the following limitations. Firstly, our study’s cross-sectional design would not allow us to determine direct causality inference. Therefore, additional research is required to verify the association between the risk factors and their impact on the outcome. Secondly, the MetS criteria used for this study were not specific for the African population, as the validated cut-off points were designed for the European, American, and Asian populations. [59,60]. Thirdly, the WHO tool was utilised to measure WC, and was subsequently used as a measurement to evaluate the prevalence of all three defined MetS criteria for this study. The WHO-recommended site for the measurement of WC is at the midpoint between the lowest rib and the superior border of the iliac crest, whereas ATP III and Harmonized ATP III measurement of WC is directly above the superior border of the iliac crest [61]. As a result, the prevalence rates reported in this review may not accurately reflect the true prevalence rate in SSA. Finally, we acknowledge that our study recruited mostly young people, which might affect our findings’ accuracy.

Despite these limitations, the study was powered to generate statistically significant results that reflect the adult population in Sierra Leone. Our study is the first to report the prevalence of MetS among adults in Freetown, Sierra Leone, using the ATP III, IDF, and Harmonized ATP III criteria.

## 5. Conclusions

The findings of the community-based survey suggest that MetS is a major public health burden in Sierra Leone, with a strong correlation to an increased risk of cardiovascular disease. To decrease the high prevalence of MetS in Sierra Leone, it is imperative to develop effective controlled strategies that necessitate tackling obesity, reducing sedentary behaviour and improving physical activities.

## Data Availability

All data produced in the present study are available upon reasonable request to the authors

## Acknowledgements

We thank everyone who participated in the study and the research assistants who helped make it possible.

## Authors’ contributions

JBWR, TRK, SKS, and SS have contributed equally to this work. JBWR, SS, and SC created the study’s design. VC and MS were in charge of acquiring data projects and recruiting participants for the community. SKS and TRK conducted the statistical analyses. JBWR, SS, AB and OTA wrote the first draft. OZM, JC, AJ, and SL reviewed and edited the final manuscript. DRL reviewed all stages of the drafted manuscript for important intellectual content. All authors contributed to the data.

